# How Should COVID-19 Vaccines be Distributed between the Global North and South? A Discrete Choice Experiment in Six European Countries

**DOI:** 10.1101/2022.05.19.22275055

**Authors:** JI Steinert, H Sternberg, GA Veltri, T Büthe

## Abstract

**Background:** The global distribution of COVID-19 vaccinations remains highly unequal. We examine public preferences in six European countries regarding the allocation of COVID-19 vaccines between the Global South and Global North.

**Methods:** We conducted online discrete choice experiments with adult participants in France (n=766), Germany (n=1964), Italy (n=767), Poland (n=670), Spain (n=925), and Sweden (n=938). Respondents were asked to decide which one of two candidates, who varied along four attributes: age, mortality risk, employment, and living in a low- or high-income country, should receive the vaccine first. We analysed the relevance of each attribute in allocation decisions using a conditional logit regression.

**Results:** Across countries, respondents selected candidates with a high mortality and infection risk, irrespective of whether the candidate lived in their own country. All else equal, respondents in Italy, France, Spain, and Sweden gave priority to a candidate from a low-income country, whereas German respondents were significantly more likely to choose the candidate from their own country. Female, younger, and more educated respondents were more favourable of an equitable vaccine distribution.

**Conclusions:** Given these preferences for global solidarity, European governments should promote vaccine transfers to poorer world regions.

**Funding:** Funding was provided by the European Union’s Horizon H2020 research and innovation programme under grant agreement 101016233 (PERISCOPE).

## Introduction

In his opening speech to address the United Nations General Assembly in September 2021, Secretary General António Guterres expressed stark discontent with the highly unequal global distribution of COVID-19 vaccines: *“A majority of the wealthier world is vaccinated. Over 90 percent of Africans are still waiting for their first dose. This is a moral indictment of the state of our world. It is an obscenity*.*”*(UN Secretary General, 2021). At the time of writing, only 10% of citizens in low-income countries have received at least one dose of a COVID-19 vaccine, while more than 80% of available doses have been allocated to people in high-income countries.(Padma, 2021) This blatant inequity in access to COVID-19 vaccines is partly a consequence of widespread vaccine nationalism in high-income countries, including the stockpiling of vaccine doses for their own citizens.(Harman et al., 2021; Herzog et al., 2021; Wagner et al., 2021)

To ensure “fair and equitable access” to COVID-19 vaccines for all countries, the World Health Organisation (WHO), the Vaccine Alliance (Gavi), and the Coalition for Epidemic Preparedness Innovations (CEPI) formed a multilateral initiative named “COVID-19 Vaccines Global Access” (COVAX).(Herzog et al., 2021) However, several governments have resorted to making bilateral purchasing agreements with vaccine manufacturers outside of COVAX, which has substantially weakened the initiatives’ collective purchasing power.(Kim, 2021; Wouters et al., 2021) Moreover, recent estimates published by “Think Global Health” suggest that only a small fraction of the promised vaccine doses have actually reached people in low-income countries. The WHO’s pledges for a more equitable COVID-19 vaccine distribution have not been fulfilled.

Ramping up global COVID-19 vaccination rates is imperative for three reasons. From an ethical perspective, unequal access to vaccination leads to high rates of transmission, severe infections, and deaths in those parts of the world where health care capacity is the lowest. This aggravates existing health inequities between the Global South and North.(Godlee, 2021; Katz et al., 2021; Krause et al., 2021; Wagner et al., 2021) In addition, achieving a more equitable vaccine distribution would have utilitarian benefits: A recent modeling study compared two hypothetical scenarios – one in which the 50 richest countries used all available vaccines and one in which vaccines were allocated to all countries proportionally to their population size – finding that the former scenario would lead to twice as many COVID-19 deaths.(Herzog et al., 2021) Second, there are compelling economic arguments in favour of equitable vaccine distribution: The RAND Corporation estimates that constrained access to COVID-19 vaccines in low-and middle-income countries (LMICs) would reduce the global GDP by US$ 153 billion each year, including a loss of US$ 56 billion in the European Union and United States combined.(Hafner et al., 2020) Put differently, every US$ 1 spent on supplying vaccines to LMICs would yield a return of US$ 4.8.(Hafner et al., 2020) Third, alleviating global asymmetries in COVID-19 vaccine coverage is warranted for virologic reasons. Unmitigated COVID-19 transmissions in some parts of the world will create evolutionary reservoirs from which new SARS-CoV-2 variants could arise, increasing the risk of immune escape - for both vaccine-induced and natural immunity - and of other phenotypic changes that could lead to greater virulence.(Saad-Roy et al., 2021; Telenti et al., 2021; van Oosterhout et al., 2021; Wagner et al., 2021) As WHO Director General Tedros Adhanom Ghebreyesus put it: “none of us will be safe until everyone is safe”.(UN Secretary General, 2021b)

Decision-makers in high-income countries are incentivised to engage in vaccine nationalism if there is limited public support for giving COVID-19 vaccines to poorer regions of the world.(Clarke et al., 2021) Public opinion can be affected by populist rhetoric from nationalist parties, portraying vaccine donations to the Global South as a lack of patriotism. Governments will likely only donate vaccines or actively participate in international vaccine alliances such as COVAX if they do not expect to pay a price at the ballot box. A thorough understanding of public preferences for the global distribution of COVID-19 vaccines is therefore paramount.

Recent empirical literature has explored public preferences for the allocation of COVID-19 vaccines. A majority of these studies examined public opinion on *prioritisation within* high-income countries and when COVID-19 vaccine availability was still heavily constrained in those countries.(Duch et al., 2021; Gollust et al., 2020; Knotz et al., 2021; Luyten et al., 2020; Persad et al., 2021; Reeskens et al., 2021; Sprengholz et al., 2021) Based on data from online surveys and survey experiments, the studies revealed substantial public support for prioritising frontline healthcare workers and clinically vulnerable groups.(Duch et al., 2021; Persad et al., 2021) To our knowledge, only four studies to date have examined individuals’ preferences on the distribution of COVID-19 vaccines across national borders.(Clarke et al., 2021; Guidry et al., 2021; Klumpp et al., 2021; Vanhuysse et al., 2021) One online survey conducted in seven high-income countries found that around 50% of participants generally supported global allocation schemes that would give priority to the countries that could not afford to purchase vaccines.(Clarke et al., 2021) Another survey conducted in the US found that 40% of respondents were in favour of donating at least 10% of the nationally purchased vaccines to poorer countries. Support was less pronounced among older respondents – a group that is at greater risk of severe disease progression if infected.(Guidry et al., 2021)

A survey conducted in Germany asked participants to choose between different options for international agreements and alliances on the distribution of COVID-19 vaccines, which varied by (i) countries joining the agreement, (ii) distribution rules, and (iii) cost per German household. The authors found that participants displayed a strong preference for an alliance exclusively composed of EU states. More importantly, the authors found that participants were more supportive of vaccine alliances if the national cost of participation was lower and national vaccine coverage higher, suggesting that participants’ preferences were significantly shaped by self-interest.(Vanhuysse et al., 2021)

In contrast, in a discrete choice experiment conducted in Germany and the US, participants in both countries expressed a strong preference for prioritising vaccine allocation to countries with a higher number of COVID-19 deaths and fewer intensive care unit beds, even when they were asked to imagine that they or a vulnerable family member were still waiting for the COVID-19 vaccine.(Klumpp et al., 2021) Notably, no previous study to date has exclusively sampled participants who were still waiting for their first COVID-19 vaccine dose when participating in the survey experiment.

In this paper, we analyse new experimental evidence from six EU countries on citizens’ preferences for the distribution of COVID-19 vaccines between the Global South and North. We advance the literature in three ways. First, by covering six countries, we implement the largest survey experiment on international vaccine allocation preferences to date and are thus able to examine differences in citizens’ preferences across EU member states. Second, we conduct a discrete choice experiment among participants who are themselves not yet vaccinated, asking them to allocate a COVID-19 vaccine to either a person in their own country or to a person in a country in the Global South. This places specific salience on the notion that donating a vaccine dose to a person in the Global South might mean sacrificing one’s own dose or that of a fellow citizen, thus leveraging self-interest-based and nationalistic considerations. Third, we specifically examine heterogeneity in participants’ preferences along key sociodemographic characteristics. Policymakers can use these insights to anticipate which population groups will be most and least supportive of COVID-19 vaccine donations.

## Results

### Sample characteristics

A total of 6,030 eligible participants across all six countries completed the DCE. We excluded participants who did not reply to all of the eight choice tasks in the DCE. To that end, attrition across choice tasks ranged from 0% in Germany and Sweden, roughly 10% in France, Poland and Spain, to almost 15% in Italy. Table 1 presents socioeconomic characteristics of participants in each country. The German sample shows higher proportions of (i) older participants (age groups 55-64 and 65+ years), (ii) less educated participants, and (iii) participants with an increased risk of a severe COVID-19 infection. Cross-country differences in the sample compositions and deviations from the census statistics are probably linked to the timing of the survey launch and the vaccination progress in each country. The German survey was launched earlier, when a higher number of older people were not yet eligible for the COVID-19 vaccination and therefore were eligible to participate in the survey.

**Table 1.**
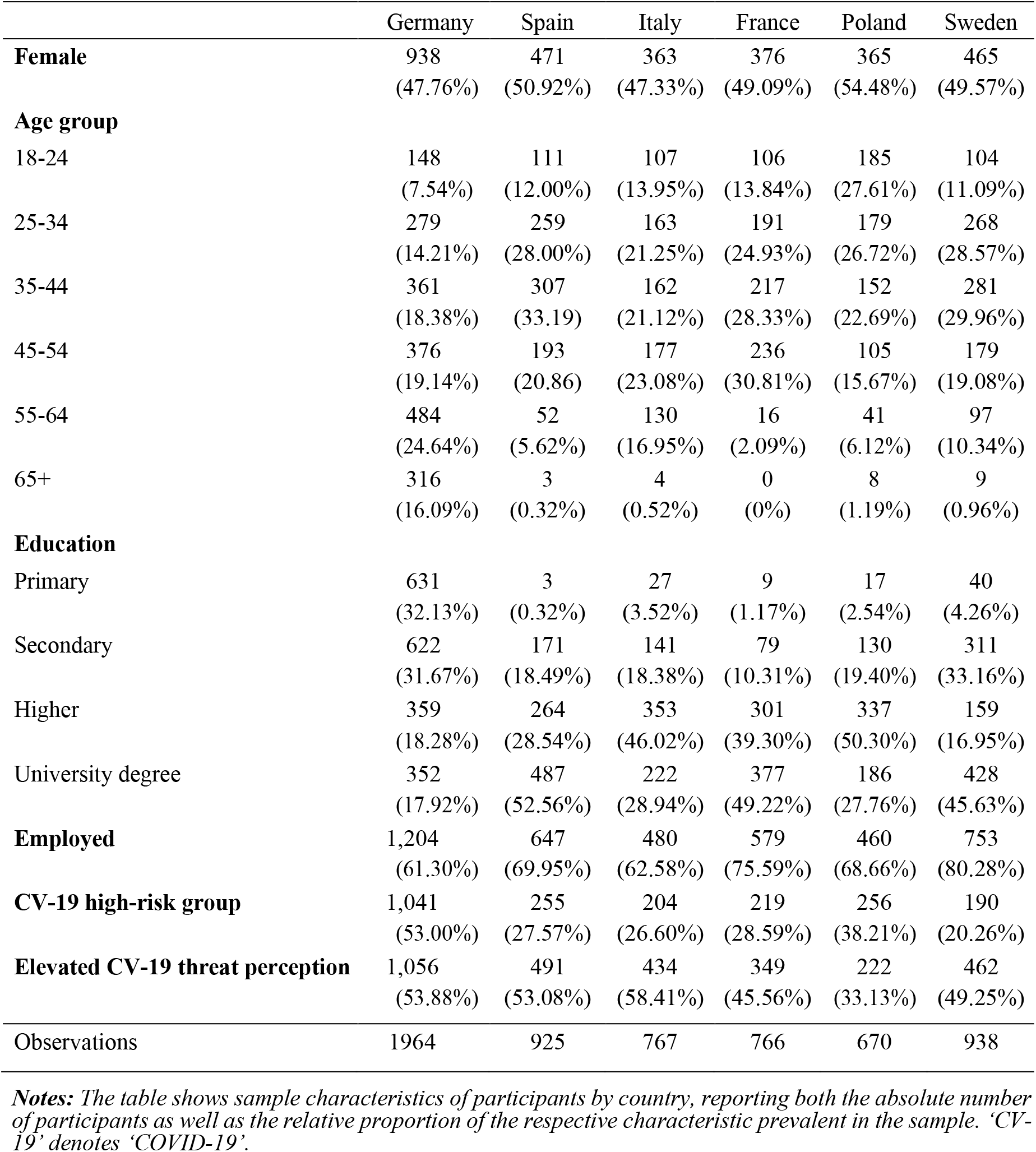
Socio-demographic characteristics

|  | Germany | Spain | Italy | France | Poland | Sweden |
| --- | --- | --- | --- | --- | --- | --- |
| **Female** | 938<br>(47.76%) | 471<br>(50.92%) | 363<br>(47.33%) | 376<br>(49.09%) | 365<br>(54.48%) | 465<br>(49.57%) |
| **Age group** |  |  |  |  |  |  |
| 18-24 | 148<br>(7.54%) | 111<br>(12.00%) | 107<br>(13.95%) | 106<br>(13.84%) | 185<br>(27.61%) | 104<br>(11.09%) |
| 25-34 | 279<br>(14.21%) | 259<br>(28.00%) | 163<br>(21.25%) | 191<br>(24.93%) | 179<br>(26.72%) | 268<br>(28.57%) |
| 35-44 | 361<br>(18.38%) | 307<br>(33.19) | 162<br>(21.12%) | 217<br>(28.33%) | 152<br>(22.69%) | 281<br>(29.96%) |
| 45-54 | 376<br>(19.14%) | 193<br>(20.86) | 177<br>(23.08%) | 236<br>(30.81%) | 105<br>(15.67%) | 179<br>(19.08%) |
| 55-64 | 484<br>(24.64%) | 52<br>(5.62%) | 130<br>(16.95%) | 16<br>(2.09%) | 41<br>(6.12%) | 97<br>(10.34%) |
| 65+ | 316<br>(16.09%) | 3<br>(0.32%) | 4<br>(0.52%) | 0<br>(0%) | 8<br>(1.19%) | 9<br>(0.96%) |
| **Education** |  |  |  |  |  |  |
| Primary | 631<br>(32.13%) | 3<br>(0.32%) | 27<br>(3.52%) | 9<br>(1.17%) | 17<br>(2.54%) | 40<br>(4.26%) |
| Secondary | 622<br>(31.67%) | 171<br>(18.49%) | 141<br>(18.38%) | 79<br>(10.31%) | 130<br>(19.40%) | 311<br>(33.16%) |
| Higher | 359<br>(18.28%) | 264<br>(28.54%) | 353<br>(46.02%) | 301<br>(39.30%) | 337<br>(50.30%) | 159<br>(16.95%) |
| University degree | 352<br>(17.92%) | 487<br>(52.56%) | 222<br>(28.94%) | 377<br>(49.22%) | 186<br>(27.76%) | 428<br>(45.63%) |
| **Employed** | 1,204<br>(61.30%) | 647<br>(69.95%) | 480<br>(62.58%) | 579<br>(75.59%) | 460<br>(68.66%) | 753<br>(80.28%) |
| **CV-19 high-risk group** | 1,041<br>(53.00%) | 255<br>(27.57%) | 204<br>(26.60%) | 219<br>(28.59%) | 256<br>(38.21%) | 190<br>(20.26%) |
| **Elevated CV-19 threat perception** | 1,056<br>(53.88%) | 491<br>(53.08%) | 434<br>(58.41%) | 349<br>(45.56%) | 222<br>(33.13%) | 462<br>(49.25%) |
| Observations | 1964 | 925 | 767 | 766 | 670 | 938 |

*Notes: The table shows sample characteristics of participants by country, reporting both the absolute number of participants as well as the relative proportion of the respective characteristic prevalent in the sample. 'CV-19' denotes 'COVID-19'.*

### Discrete Choice Experiment

Table 2 summarises results for the main effects model, which shows, separately for each country, the impacts of the four attributes and levels on a candidate’s likelihood of being chosen by the respondent to receive the COVID-19 vaccine first. In France, Italy, Spain, and Sweden, respondents, on average, chose the hypothetical candidate from the Global South over the hypothetical candidate from their own country to receive the vaccine first (Spain: OR: 1.79, 95% CI: 1.55-2.06; Italy: OR: 1.74, 95% CI: 1.50-2.01; Sweden: OR: 1.43, 95% CI: 1.24-1.65;

**Table 2.** Main attribute effects by country

| **Outcome:** Choosing the respective candidate to receive the vaccine |  |  |  |  |  |  |
| --- | --- | --- | --- | --- | --- | --- |
|  | Germany<br>(1) | Spain<br>(2) | Italy<br>(3) | France<br>(4) | Poland<br>(5) | Sweden<br>(6) |
| **Age** |  |  |  |  |  |  |
| 20 years | Reference category |  |  |  |  |  |
| 40 years | 1.39***<br>[1.33,1.45] | 1.25***<br>[1.17,1.33] | 1.16***<br>[1.09,1.22] | 1.15***<br>[1.08,1.22] | 1.08*<br>[1.02,1.14] | 1.13***<br>[1.06,1.20] |
| 60 years | 0.87***<br>[0.83,0.90] | 0.85***<br>[0.80,0.91] | 0.88***<br>[0.84,0.94] | 0.87***<br>[0.82,0.93] | 0.85***<br>[0.80,0.91] | 0.77***<br>[0.73,0.82] |
| 80 years | 0.81***<br>[0.73,0.90] | 1.23**<br>[1.06,1.43] | 1.12<br>[0.94,1.33] | 0.59***<br>[0.50,0.69] | 0.94<br>[0.78,1.13] | 0.70***<br>[0.61,0.81] |
| **COVID-19 mortality risk** |  |  |  |  |  |  |
| Average | Reference category |  |  |  |  |  |
| Increased | 5.63***<br>[5.13,6.19] | 3.05***<br>[2.66,3.50] | 2.27***<br>[1.96,2.64] | 4.54***<br>[3.88,5.31] | 2.40***<br>[2.07,2.79] | 5.67***<br>[4.91,6.56] |
| Strongly increased | 13.23***<br>[11.64,15.03] | 9.68***<br>[8.12,11.53] | 4.36***<br>[3.69,5.15] | 9.04***<br>[7.50,10.91] | 4.35***<br>[3.65,5.19] | 14.20***<br>[11.75,17.17] |
| **Employment situation** |  |  |  |  |  |  |
| Not employed | Reference category |  |  |  |  |  |
| Employed (guaranteed income) | 1.30***<br>[1.18,1.42] | 0.79**<br>[0.68,0.92] | 1.14<br>[0.99,1.32] | 1.04<br>[0.89,1.22] | 1.06<br>[0.91,1.24] | 1.00<br>[0.87,1.16] |
| Employed (income losses) | 2.37***<br>[2.19,2.56] | 1.91***<br>[1.71,2.13] | 1.67***<br>[1.49,1.87] | 1.65***<br>[1.47,1.86] | 1.36***<br>[1.21,1.53] | 1.73***<br>[1.54,1.94] |
| Essential services | 6.01***<br>[5.38,6.72] | 4.05***<br>[3.50,4.70] | 2.27***<br>[1.98,2.59] | 2.74***<br>[2.33,3.22] | 1.88***<br>[1.65,2.15] | 4.22***<br>[3.57,4.99] |
| **Country of residence** |  |  |  |  |  |  |
| Respondents' country | Reference category |  |  |  |  |  |
| Global South | 0.69***<br>[0.62,0.76] | 1.79***<br>[1.55,2.06] | 1.74***<br>[1.50,2.01] | 1.37***<br>[1.18,1.59] | 0.99<br>[0.86,1.15] | 1.43***<br>[1.24,1.65] |
| Log likelihood | -14428.91 | -7090.45 | -6541.07 | -5994.74 | -5860.07 | -6875.04 |
| AIC | 28875.82 | 14198.89 | 13100.14 | 12007.48 | 11738.15 | 13768.07 |
| BIC | 28951.02 | 14267.31 | 13166.88 | 12074.21 | 11803.67 | 13836.62 |
| Pseudo $R^2$ | 0.22 | 0.19 | 0.10 | 0.17 | 0.08 | 0.23 |
| Observations | 31424 | 14800 | 12272 | 12256 | 10720 | 15008 |

**Notes:** Coefficients are odds ratios based on conditional logit estimations with standard errors clustered at the individual level. Results to be interpreted relative to the indicated reference category. 95% confidence intervals in brackets. \* $p < 0.05$ , \*\* $p < 0.01$ , \*\*\* $p < 0.001$ . Sample sizes reflect the eight choice tasks performed by each respondent.

France: OR: 1.37, 95% CI: 1.18-1.59; all p-values<0.001). For German respondents, we observe the opposite pattern: candidates from the Global South had significantly lower odds of being chosen to receive the vaccine (OR: 0.69, 95% CI: 0.62-0.76, p-value<0.001). In Poland, a candidate’s country of residence neither increased nor decreased the odds of being chosen to receive the vaccine (OR: 0.99, 95% CI: 0.86-1.15).

For the attribute of COVID-19 mortality risk, we observe a similar pattern in all surveyed countries: The odds of being chosen to receive the vaccine were between two and almost six times higher for a candidate with an increased COVID-19 mortality risk, relative to a candidate with an average risk. The effect was even more pronounced for a candidate with a strongly increased mortality risk (ranging from OR: 4.35, 95% CI: 3.65-5.19 in Poland to OR: 14.20, 95% CI: 11.75-17.17 in Sweden; all p-values<0.001).

For employment status and age, we also observed largely similar patterns across countries: First, employed candidates who lost income due to the pandemic and candidates who were employed in essential services exhibited were significantly more likely to be chosen to receive the vaccine when compared to unemployed candidates. Second, in all countries, 40-year-old candidates had slightly higher odds of being chosen to receive the vaccine than 20-or 60-year-old candidates.

Figure 1 illustrates the results of the heterogeneity analysis that was pooled across countries (see Tables S2-3 for exact coefficients by subgroup and interaction terms). The odds of choosing the candidate from the Global South rather than the candidate from the respondents’ own country were significantly higher for female (OR of interaction: 1.22, 95% CI: 1.10-1.36, p-value<0.001) and more educated (OR of interaction: 1.63, 95% CI: 1.46-1.82, p-value<0.001) respondents, and significantly lower for older respondents (above 45 years) (OR of interaction: 0.68, 95% CI: 0.61-0.76, p-value<0.001). In contrast, respondents who were themselves at high risk of a severe COVID-19 disease progression were significantly less supportive of distributing the COVID-19 vaccine to a candidate from the Global South (OR of interaction: 0.70, 95% CI: 0.63-0.78, p-value<0.001). In the subgroup of employed respondents, the odds of distributing the vaccine to a candidate in the Global South were slightly lower than in the subgroup of unemployed respondents (OR of interaction: 0.89, 95% CI: 0.80-1,00, p-value=0.045). The degree of COVID-19 threat perception did not seem to significantly affect respondents’ distribution preferences (OR of interaction: 1.10, 95% CI: 0.99-1.22, p-value=0.071).

**Figure 1.**
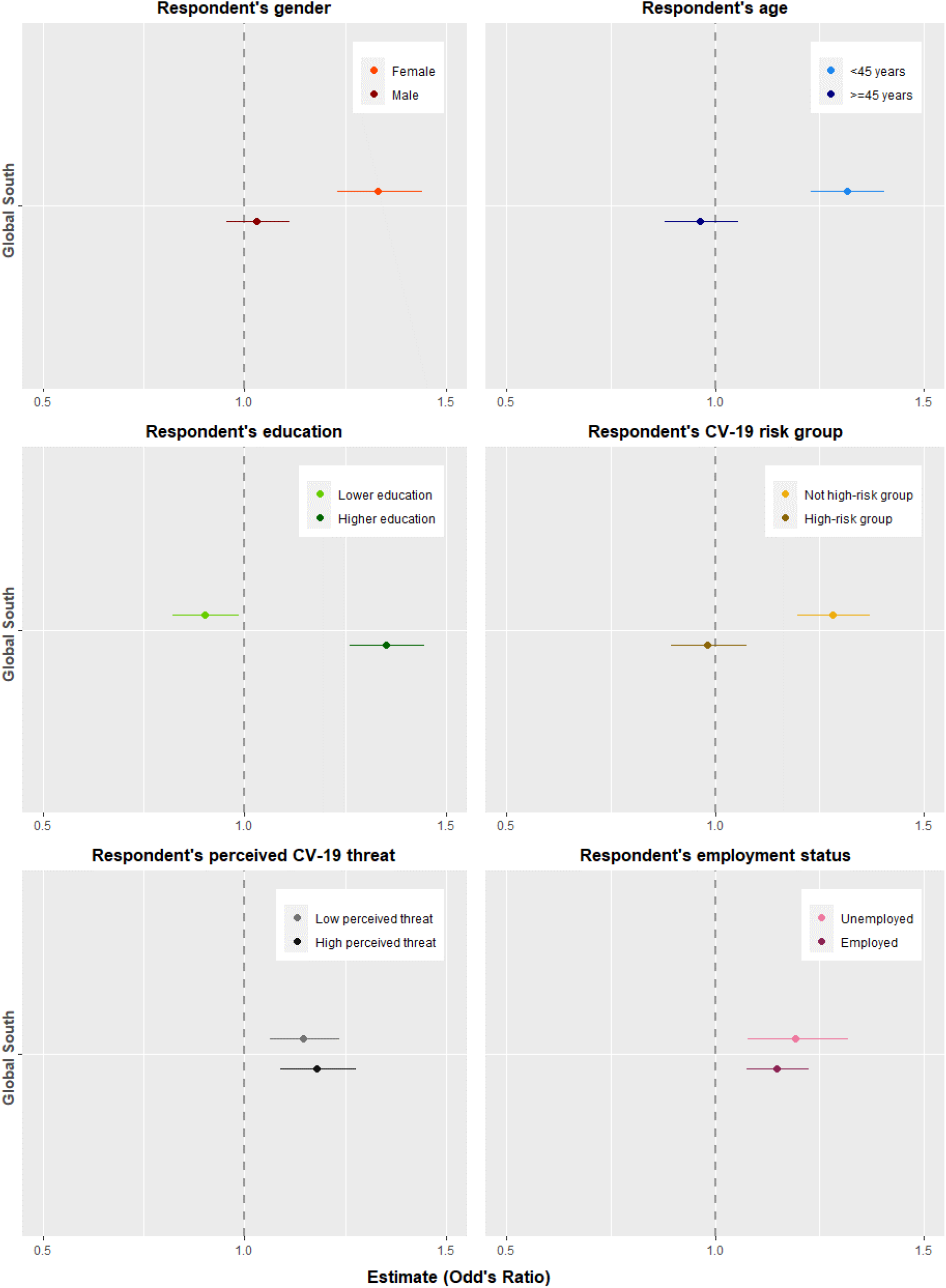
Heterogeneity of country attribute by respondent characteristics (pooled sample)

Figure S3 and Tables S4-9 summarise the extent to which these patterns prevail in each country. The heterogeneities we identify were strongest in the German, Spanish, and Swedish samples. In the French and Italian sample, these patterns were prevalent, too, but not statistically significant. Interestingly, in terms of age, we found that older participants were relatively more supportive of distributing the vaccine to a candidate from the Global South in both countries. In the Italian sample, this effect was even statistically significant (OR of interaction: 1.34, 95% CI: 1.00-1.78, p-value=0.049). In the Polish survey, heterogeneity patterns were less clear, and we observed an opposite effect for educational attainment, with less educated respondents showing more pronounced preferences for an equitable vaccine distribution (OR of interaction: 0.72, 95% CI: 0.52-0.99, p-value=0.044).

## Discussion

In our DCE conducted online in six EU countries, we found widespread global solidarity and support for a more equitable distribution of COVID-19 vaccines between the Global North and South. Our results point to vaccine allocation preferences that are largely driven by the assessed vulnerability of possible recipients, irrespective of whether the recipient lives in the respondents’ own country or in the Global South. Similar to their prioritisation preferences *within* a country, our respondents – who are themselves not yet vaccinated against COVID-19 but do favour vaccination – appear to rely upon the same solidarity considerations when vaccines are distributed *between* countries. For example, they choose the candidate with the higher mortality risk over the candidate with the same nationality. In addition, we find that in a situation of vaccine scarcity and all else being equal, respondents from Spain, Italy, France and Sweden would prefer to allocate the vaccine to a person living in a country in the Global South with a worse health care system, as opposed to a person living in their own country of residence. We thus confirm the findings of studies of citizens’ distributional preferences, which revealed largely positive attitudes towards vaccine donations to poorer countries in the context of other pandemics(Kumar et al., 2012; Ritvo et al., 2013) and showed in experimental games that individuals follow egalitarian motives in their own decisions,(Dawes et al., 2007) and are even willing to punish third parties for inegalitarian behaviour.(Fehr and Gächter, 2002) Yet, we go beyond previous studies by showing that such support for allocating vaccines to low-income countries with low healthcare system capacity holds across European countries from Spain to Sweden, even among respondents who had themselves not yet gotten vaccinated, and in the context of a pandemic with truly global reach, which by the time when we conducted our experiments had lasted for well more than a year.

At the same time, our findings were inconclusive for Polish respondents and diverged for the German DCE: German survey participants were significantly less willing to allocate the COVID-19 vaccine to a person living in the Global South than to a person living in their own country, thus revealing preferences consistent with vaccine nationalism.

There are several possible explanations for the strikingly contrasting findings in Germany. First, studies have repeatedly found in-group bias (or here: national bias) in distributional preferences.(Bernhard et al., 2006; Reese et al., 2012; Yudkin et al., 2016) Such in-group bias might explain why German respondents appear willing to allocate the COVID-19 vaccines to more vulnerable candidates (e.g., those with higher COVID-19 risks) within their own country but not to candidates living in the Global South, if German participants had more nationalistic preferences, lower altruism, or less pro-social attitudes than participants from the other five countries. We lack the data to test this possible explanation directly, but it seem unlikely: several cross-country analyses investigating nationalism, patriotism, and xenophobia have found German citizens to be among the least nationalistic in Europe.(Coenders et al., 2021; Lubbers and Coenders, 2017) German citizens also do not generally appear to be outliers among their European peers in terms of altruism or similar pro-sociality variables.(Fehr et al., 2021)

Second, respondents might have been influenced by heterogeneities in the pandemic situation across countries. In Germany, our experiment was implemented as the third pandemic wave peaked, whereas in the other countries, our DCEs were launched two months later, during a phase of relatively low daily case numbers. German respondents might therefore have felt a greater urgency about having faster access to vaccination – though our data does not show evidence of a substantially higher reported COVID-19 threat perception among German respondents (see Table S10 and Figure S4).

Third, the findings could be a function of the differences in the timing of the surveys relative to the progress of national vaccination campaigns. A recent survey experiment found that individuals with a higher perceived rank in the global income distribution feel stronger pressure to donate.(Fehr et al., 2021) Extrapolating this dynamic to the context of our study, we might expect to see a higher individual inclination for vaccine donations in countries where the (perceived) vaccination rate is high by international standards. When the survey was fielded in Italy, France, Spain, Poland, and Sweden, the vaccination rate in those countries exceeded 50% (30%) for first (second) doses whereas in Germany it was only at 25% (8%) (see Table S9). This difference in domestic conditions may have affected the perceived scarcity of the COVID-19 vaccines between countries and the normative assessment thereof.

A number of limitations are worth noting. First, participants’ preferences may partly be driven by cross-country variation in (1) COVID-19 infection and mortality rates, (2) COVID-19 vaccination rates, and (3) the pandemic trajectory over time. Our analyses did not allow for any in-depth investigation of these factors. Second, on an individual level, there may be additional characteristics that explain approval or rejection of COVID-19 vaccine donations but were not captured in the survey, including (1) nationalistic attitudes, (2) altruistic preferences or (3) migration background. Third, our analysis points to a number of predictors of variation.

However, they are not susceptible to experimental manipulation and should therefore not be interpreted as causal. Fourth, at the country level (except in Germany), we were not able to achieve our initial target sample sizes due to budget constraints. Thus, we likely only have adequate statistical power in the pooled analysis. Lastly and relatedly, sample sizes varied across countries and statistical power was higher in the German survey than in the other five surveys. However, since we find statistically significant effects of the main attributes in all countries except from Poland, lack of statistical power might have been less of a concern.

Policymakers and global health scholars have condemned the unequitable distribution of COVID-19 vaccines between high- and low-income countries as “vaccine apartheid”.(Gonsalves and Yamey, 2021; Harman et al., 2021) Still, the World Health Organisation’s call for a moratorium on COVID-19 booster vaccinations in high-income countries in favour of prioritising first dose vaccinations in low-income countries went unheeded, at least in part out of a sense that donating vaccines to countries in the Global South lacks popular support and might even subject the government to electoral punishment.(Krause et al., 2021) The emergence of the new Omicron variant – and fears of potential future variants – emphasises once again the transboundary nature of the COVID-19 pandemic.

Acknowledging this, governments in high-income countries should discard mitigation strategies that are guided by the premise of vaccine nationalism.(Vanhuysse et al., 2021) Findings from our study suggest that governments of European countries can rely on solid public approval for a more equitable vaccine distribution – especially among female, younger and more educated citizens. Public support for vaccine donations to the Global South may further increase once the specific COVID-19 risk groups have received their booster vaccination. More effective international policy initiatives to ensure efficient, adequate, and timely COVID-19 vaccine transfers to low-income countries are urgently needed.

## Materials and Methods

### Study Sample

We conducted an online survey experiment in six EU countries: France, Germany, Italy, Poland, Spain, and Sweden. The German survey was launched at the height of the third wave in April 2021; the other five surveys were carried out in June 2021, coinciding with a phase of low case numbers in each country (see Figure S1). In each country, we recruited respondents aged 18 years and older, drawing on online panels of the survey provider *Bilendi-Respondi*. We sampled participants based on quotas that were matched to the census population of each target country in terms of (1) gender, (2) age, (3) education, and (4) geographic location (e.g., state or province within each country) (see Table S1 for the census statistics of the sampled countries). Participants were given an individual link to the survey, where they first received information about the study’s purpose, data protection regulations, and voluntary participation. After completing the survey, participants received a voucher worth three to five Euros, which was distributed by the survey company.

### Survey Experiment

Discrete choice experiments (DCEs) are used to measure the relative importance of different characteristics that respondents weigh against each other when making certain choices.(Mangham et al., 2009) DCEs have emerged from the theoretical tradition of random utility theory and are based on the assumption that respondents express their preferences by choosing the alternative associated with the highest individual benefits or utility.(Hall et al., 2004) DCEs have advantages over other stated preference techniques, such as ranking or rating exercises, because they (i) more closely mimic real-world choice scenarios, (ii) reduce the cognitive complexity for respondents, and (iii) can elicit implicit preferences.(Mangham et al., 2009)

We used the DCE methodology to elicit participants’ preferences on the allocation of a COVID-19 vaccine dose to a person in the Global North or to a person in the Global South. We presented eight different choice sets and asked respondents to choose whether Person A or Person B should receive the COVID-19 vaccine first. Respondents were told that the other candidate of each pair would have to wait substantially longer to receive their first vaccine dose. In each of the eight choice sets, one candidate was described as living in the country of residence of the respondent – i.e. a high-income country with high healthcare system capacity, and the other candidate was described as living in a low-income country with a low healthcare system capacity. The healthcare system capacity was explicitly mentioned in the two candidate profiles. Across choice sets, candidates’ characteristics varied along three additional attributes, namely (1) age (20; 40; 60; 80 years), (2) individual COVID-19 mortality risk due to comorbidity and/or lifestyle (no increased risk; increased risk; strongly increased risk), and (3) employment status (not employed; employed and guaranteed income; employed and income losses due to COVID-19 restrictions; employed in essential services).

The specific combination of candidate profiles in the eight choice sets, i.e., the experimental design (presented in Table 4) was selected for statistical efficiency (referred to as “D-efficiency”), which is accomplished by minimising the asymptotic variance-covariance matrix of the parameter estimates (based on an algorithm implemented in the software Ngene).(Reed Johnson et al., 2013) A sample choice situation as presented to respondents in the online survey, along with further technical information, is shown in Figure S2 and notes.

**Table 4.** Full list of choice sets of the DCE

| Choice set | Age in years | COVID-19 mortality risk | Employment status | Country of residence and healthcare system capacity |
| --- | --- | --- | --- | --- |
| 1<br>Person A | 40 | No increased risk due to comorbidity and/or lifestyle | Not employed | [Respondents' country of residence], with high healthcare system capacity |
| 1<br>Person B | 40 | Strongly increased risk due to comorbidity and/or lifestyle | Employed and guaranteed income | Low-income country, with poor healthcare system capacity) |
| 2<br>Person A | 60 | Strongly increased risk due to comorbidity and/or lifestyle | Employed and guaranteed income | [Respondents' country of residence], with high healthcare system capacity |
| 2<br>Person B | 60 | No increased risk due to comorbidity and/or lifestyle | Employed in essential services | Low-income country, with poor healthcare system capacity) |
| 3<br>Person A | 60 | Increased risk due to comorbidity and/or lifestyle | Employed and income losses due to COVID-19 restrictions | [Respondents' country of residence], with high healthcare system capacity |
| 3<br>Person B | 80 | Increased risk due to comorbidity and/or lifestyle | Not employed | Low-income country, with poor healthcare system capacity |
| 4<br>Person A | 20 | Increased risk due to comorbidity and/or lifestyle | Employed in essential services | Low-income country, with poor healthcare system capacity |
| 4<br>Person B | 40 | No increased risk due to comorbidity and/or lifestyle | Employed and income losses due to COVID-19 restrictions | [Respondents' country of residence], with high healthcare system capacity |
| 5<br>Person A | 40 | No increased risk due to comorbidity and/or lifestyle | Employed in essential services | [Respondents' country of residence], with high healthcare system capacity |
| 5<br>Person B | 20 | Increased risk due to comorbidity and/or lifestyle | Not employed | Low-income country, with poor healthcare system capacity |
| 6<br>Person A | 20 | Strongly increased risk due to comorbidity and/or lifestyle | Employed and income losses due to COVID-19 restrictions | Low-income country, with poor healthcare system capacity |
| 6<br>Person B | 20 | No increased risk due to comorbidity and/or lifestyle | Employed and guaranteed income | [Respondents' country of residence], with high healthcare system capacity |
| 7<br>Person A | 80 | Increased risk due to comorbidity and/or lifestyle | Not employed | Low-income country, with poor healthcare system capacity |
| 7<br>Person B | 60 | Increased risk due to comorbidity and/or lifestyle | Employed and income losses due to COVID-19 restrictions | [Respondents' country of residence], with high healthcare system capacity |
| 8<br>Person A | 40 | No increased risk due to comorbidity and/or lifestyle | Employed and guaranteed income | Low-income country, with poor healthcare system capacity |
| 8<br>Person B | 40 | Strongly increased risk due to comorbidity and/or lifestyle | Employed in essential services | [Respondents' country of residence], with high healthcare system capacity |

**Notes:** [Respondents' country of residence] was differed depending on the respective country the survey was fielded in, e.g., in the German sample, this attribute level was 'Germany, with high healthcare system capacity'. The design was determined with a built-in constraint for the attributes Age and Employment status in order to avoid implausible combinations (specifically, an age of 80 was always combined with not being employed).

Participants were only eligible to participate in the survey experiment if they (1) had not yet been vaccinated against COVID-19 at the time of the survey and (2) indicated that they were willing to get vaccinated. We employed these two criteria to elicit prioritisation preferences on how to distribute scarce COVID-19 vaccine doses among individuals who perceived the vaccine as beneficial *and* were themselves still waiting to receive their first dose. Thus, the decisions of participants in the survey experiment might to a larger extent be informed by the fear/worry of having to sacrifice their own chances of getting a vaccine.

Power calculations for the discrete choice experiment indicated a desired sample size of n=2,061 respondents per country.(de Bekker-Grob et al., 2015) However, the rapidly evolving pandemic situation in combination with conditional eligibility on vaccination status (see above) only allowed us to reach the required sample size in Germany due to the earlier timing of the data collection. Sample sizes in the remaining countries ranged from 670 in Poland to 925 in Spain, resulting in a total sample size of 6,030 eligible responses.

### Heterogeneity Variables

Similar to previous studies,(Duch et al., 2021; Persad et al., 2021) we examined the effect of the attribute ‘Country of residence’ for heterogeneity in terms of respondents’ socioeconomic characteristics. Specifically, we assessed whether respondents’ (i) gender, (ii) age, (iii) education, (iv) individual COVID-19 risk status, (v) COVID-19 threat perception, and (vi) employment status predicted differences in their allocation choices. Considering the limited statistical power within countries, the heterogeneity analysis was conducted with the pooled sample.

### Statistical Analyses

The empirical analysis comprised three steps. First, we estimated the main effects model for each country separately to assess the impact of the four candidate attributes (country of residence, age, COVID-19 mortality risk, employment status) on the probability of choosing a specific candidate. For each of the six countries, we estimated a conditional logit model by regressing the respondents’ allocation choice on the attribute levels of the candidate. Second, we examined heterogeneity in the effect of the country of residence attribute by adding interaction terms between country of residence and heterogeneity variables to the above regression. A pre-analysis plan along with justifications of any deviations thereof is accessible via https://osf.io/72jrq/.

### Ethical Approval

The study received approvals from the ethics committees of the medical faculty at the Technical University of Munich (TUM, IRB 227/20 S) and the ethics board at the University of Trento (Trento, IRB 2021-027).

## Data Availability

All data and code are available online at https://osf.io/72jrq/

## Acknowledgements

We thank everyone who helped with translating: Walter Osika, Jocelyn Raude, Jonathan Garcia Fuentes, Anna Glyk, and Kathrin and Michal Bartoszewski. This project was funded by the European Union’s Horizon 2020 research and innovation programme under grant agreement No 101016233 PERISCOPE). We are grateful for helpful comments on the analysis and interpretation made by Philipp Lergetporer, Michael Kurschilgen and Abu Siddique.

## Author contributions

GAV, TB, and JIS acquired funding for this research study. JIS, HS and TB conceptualised and led the study and developed the pre-analysis plan. JIS, GAV and HS oversaw and managed the data collection. JIS and HS merged and cleaned the data, and HS conducted the data analyses and data visualisations. JIS, HS, TB, and GAV contributed to the interpretation of the quantitative findings. JIS and HS drafted the first version of the manuscript and all authors provided substantial revisions and feedback. We confirm that all authors have read and approved the final version of the manuscript.

## Data availability statement

A pre-analysis plan along with justifications of any deviations thereof is accessible via the Open Science Framework website: https://osf.io/72jrq/. All data and code will be shared via the same website.

