## Supplementary Files for "How Should COVID-19 Vaccines be Distributed between the Global North and South? A Discrete Choice Experiment in Six European Countries"

### Tables

**Table S1 -** Population Census Data for Key Sociodemographic Variables

| Country | female | 18-24 years | 25-34 years | 35-44 years | 45-54 years | 55-64 years | 65+ years | low education | medium education | high education |
|---|---|---|---|---|---|---|---|---|---|---|
| France | 51.67% | 8.09% | 11.6% | 12.4% | 13.3% | 12.6% | 20.4% | 22.3% | 42.4% | 35.3% |
| Germany | 50.66% | 7.41% | 12.7% | 12.3% | 14.4% | 14.9% | 21.8% | 20.1% | 52.7% | 27.2% |
| Italy | 51.29% | 6.91% | 10.7% | 13.0% | 16.1% | 14.1% | 23.2% | 39.0% | 43.1% | 17.9% |
| Poland | 51.60% | 7.26% | 14.2% | 16.1% | 12.6% | 13.4% | 18.2% | 12.9% | 58.1% | 28.9% |
| Spain | 50.99% | 7.54% | 10.2% | 15.4% | 15.9% | 13.3% | 19.6% | 38.2% | 25.8% | 36.0% |
| Sweden | 49.69% | 7.89% | 14.1% | 12.4% | 13.0% | 11.5% | 20.0% | 20.7% | 41.1% | 38.3% |

**Table S2** – Country of residence attribute main effect - By subgroups of respondent characteristics (pooled sample)

| | Full sample | Male | Female | <45 yrs. | >45 yrs. | Low edu | High edu | Not high-risk | High-risk | Low threat | High threat | Unemployed | Employed |
|---|---|---|---|---|---|---|---|---|---|---|---|---|---|
| | (1) | (2) | (3) | (4) | (5) | (6) | (7) | (8) | (9) | (10) | (11) | (12) | (13) |
| **Country of residence** | | | | | | | | | | | | | |
| Respondents' country | Reference category | | | | | | | | | | | | |
| Global South | 1.16*** | 1.03 | 1.33*** | 1.32*** | 0.96 | 0.90* | 1.35*** | 1.28*** | 0.98 | 1.15*** | 1.18*** | 1.19*** | 1.15*** |
| | [1.10,1.23] | [0.96,1.11] | [1.23,1.44] | [1.23,1.41] | [0.88,1.06] | [0.82,0.99] | [1.26,1.44] | [1.20,1.37] | [0.89,1.07] | [1.06,1.24] | [1.09,1.28] | [1.08,1.32] | [1.08,1.22] |
| Pseudo $R^2$ | 0.16 | 0.13 | 0.20 | 0.16 | 0.17 | 0.15 | 0.17 | 0.17 | 0.16 | 0.14 | 0.18 | 0.18 | 0.16 |
| Observations | 96480 | 48448 | 47648 | 57280 | 38816 | 34896 | 61200 | 61456 | 34640 | 47808 | 48224 | 30128 | 65968 |

**Notes:** Outcome: Choosing the respective candidate to receive the vaccine. Coefficients are Odd's ratios based on conditional logit estimations with standard errors clustered at the individual level. Estimations were conducted with all four attributes, but only the results for the country of residence attribute is shown here. Columns 2-14 represent the exact coefficients shown in Figure 1 in the main body of the paper. Results to be interpreted relative to the indicated reference category, i.e., in the case of country of residence, relative to the preference for the vaccine being given to a person living in the country of the survey respondent answering the question. 95% confidence intervals in brackets. * $p < 0.05$, ** $p < 0.01$, *** $p < 0.001$.

**Table S3** – Country of residence attribute: Heterogeneity by respondent's characteristics (pooled results)

| | (1) | (2) | (3) | (4) | (5) | (6) | (7) |
|---|---|---|---|---|---|---|---|
| ***Country of Residence*** | | | | | | | |
| Respondents' Country | | | | Reference Category | | | |
| Global South | 1.16*** | 1.05 | 1.36*** | 0.85*** | 1.32*** | 1.11** | 1.26*** |
| | [1.10,1.23] | [0.97,1.14] | [1.27,1.45] | [0.78,0.93] | [1.24,1.41] | [1.03,1.19] | [1.14,1.38] |
| Global South × Female respondent | | 1.22*** | | | | | |
| | | [1.10,1.36] | | | | | |
| Global South × Respondent ≥ 45 | | | 0.68*** | | | | |
| | | | [0.61,0.76] | | | | |
| Global South × Higher educated respondent | | | | 1.63*** | | | |
| | | | | [1.46,1.82] | | | |
| Global South × High-risk respondent | | | | | 0.70*** | | |
| | | | | | [0.63,0.78] | | |
| Global South × High perceived threat respondent | | | | | | 1.10 | |
| | | | | | | [0.99,1.22] | |
| Global South × Employed respondent | | | | | | | 0.89* |
| | | | | | | | [0.80,1.00] |
| Log likelihood | -47779.22 | -47551.104 | -47485.96 | -47434.20 | -47501.49 | -47538.99 | -47569.78 |
| AIC | 95576.44 | 95122.21 | 94991.91 | 94888.40 | 95022.98 | 95097.99 | 95159.56 |
| BIC | 95661.73 | 95216.94 | 95086.64 | 94983.13 | 95117.71 | 95192.71 | 95254.29 |
| Pseudo $R^2$ | 0.16 | 0.16 | 0.16 | 0.17 | 0.16 | 0.16 | 0.16 |
| **Observations** | **96480** | **96096** | **96096** | **96096** | **96096** | **96032** | **96096** |

**Notes:** Outcome: Choosing the respective candidate to receive the vaccine. Coefficients are Odd's ratios based on conditional logit estimations with standard errors clustered at the individual level. Estimations were conducted with controlling for the main effects of the other three attributes, but only the results for the country of residence attribute are shown here. Columns 2-7 indicate the degree of statistical (in-)significance of the subgroup differences presented in Figure 1 in the main body of the paper and Table A1 of the supplementary material. Results to be interpreted relative to the indicated reference category, i.e., in the case of country of residence, relative to the preference for the vaccine being given to a person living in the country of the survey respondent answering the question. 95% confidence intervals in brackets. * $p < 0.05$, ** $p < 0.01$, ***$p < 0.001$.

**Table S4** – Country of residence attribute: Heterogeneity by respondent's characteristics (German sample)

| | (1) | (2) | (3) | (4) | (5) | (6) | (7) |
|---|---|---|---|---|---|---|---|
| ***Country of Residence*** | | | | | | | |
| Respondents' Country | | | | Reference Category | | | |
| Global South | 0.69*** | 0.65*** | 0.89 | 0.57*** | 0.82** | 0.70*** | 0.79** |
| | [0.62,0.76] | [0.56,0.75] | [0.77,1.03] | [0.50,0.65] | [0.71,0.94] | [0.61,0.82] | [0.67,0.92] |
| Global South × Female respondent | | 1.15 | | | | | |
| | | [0.95,1.40] | | | | | |
| Global South × Respondent ≥ 45 | | | 0.66*** | | | | |
| | | | [0.54,0.80] | | | | |
| Global South × Higher educated respondent | | | | 1.70*** | | | |
| | | | | [1.39,2.08] | | | |
| Global South × High-risk respondent | | | | | 0.73** | | |
| | | | | | [0.60,0.88] | | |
| Global South × High perceived threat respondent | | | | | | 0.96 | |
| | | | | | | [0.79,1.17] | |
| Global South × Employed respondent | | | | | | | 0.81* |
| | | | | | | | [0.66,0.99] |
| Pseudo $R^2$ | 0.22 | 0.22 | 0.23 | 0.23 | 0.22 | 0.22 | 0.22 |
| Observations | 31424 | 31424 | 31424 | 31424 | 31424 | 31424 | 31424 |

**Notes:** Outcome: Choosing the respective candidate to receive the vaccine. Coefficients are Odd's ratios based on conditional logit estimations with standard errors clustered at the individual level. Estimations were conducted with controlling for the main effects of the other three attributes, but only the results for the country of residence attribute are shown here. Columns 2-7 indicate the degree of statistical (in-)significance of the subgroup differences presented in Figure 1 in the main body of the paper and Table A1 of the supplementary material. Results to be interpreted relative to the indicated reference category, i.e., in the case of country of residence, relative to the preference for the vaccine being given to a person living in the country of the survey respondent answering the question. 95% confidence intervals in brackets. * $p < 0.05$, ** $p < 0.01$, *** $p < 0.001$.

**Table S5** – Country of residence attribute: Heterogeneity by respondent's characteristics (Spanish sample)

| | (1) | (2) | (3) | (4) | (5) | (6) | (7) |
|---|---|---|---|---|---|---|---|
| ***Country of Residence*** | | | | | | | |
| Respondents' Country | Reference Category | | | | | | |
| Global South | 1.79*** | 1.44*** | 1.83*** | 1.49* | 1.79*** | 1.47*** | 2.46*** |
| | [1.55,2.06] | [1.17,1.76] | [1.57,2.15] | [1.09,2.04] | [1.52,2.10] | [1.20,1.80] | [1.94,3.13] |
| Global South × Female respondent | | 1.54** | | | | | |
| | | [1.18,2.01] | | | | | |
| Global South × Respondent ≥ 45 | | | 0.91 | | | | |
| | | | [0.66,1.26] | | | | |
| Global South × Higher educated respondent | | | | 1.25 | | | |
| | | | | [0.89,1.76] | | | |
| Global South × High-risk respondent | | | | | 1.00 | | |
| | | | | | [0.74,1.37] | | |
| Global South × High perceived threat respondent | | | | | | 1.45** | |
| | | | | | | [1.11,1.90] | |
| Global South × Employed respondent | | | | | | | 0.63** |
| | | | | | | | [0.48,0.84] |
| Pseudo $R^2$ | 0.19 | 0.19 | 0.19 | 0.19 | 0.19 | 0.19 | 0.19 |
| Observations | 14800 | 14800 | 14800 | 14800 | 14800 | 14800 | 14800 |

**Notes:** Outcome: Choosing the respective candidate to receive the vaccine. Coefficients are Odd's ratios based on conditional logit estimations with standard errors clustered at the individual level. Estimations were conducted with controlling for the main effects of the other three attributes, but only the results for the country of residence attribute are shown here. Columns 2-7 indicate the degree of statistical (in-)significance of the subgroup differences presented in Figure 1 in the main body of the paper and Table A1 of the supplementary material. Results to be interpreted relative to the indicated reference category, i.e., in the case of country of residence, relative to the preference for the vaccine being given to a person living in the country of the survey respondent answering the question. 95% confidence intervals in brackets. * $p < 0.05$, ** $p < 0.01$, *** $p < 0.001$.

**Table S6** – Country of residence attribute: Heterogeneity by respondent's characteristics (Italian sample)

| | (1) | (2) | (3) | (4) | (5) | (6) | (7) |
|---|---|---|---|---|---|---|---|
| ***Country of Residence*** | | | | | | | |
| Respondents' Country | | | | Reference Category | | | |
| Global South | 1.74*** | 1.56*** | 1.54*** | 1.63** | 1.66*** | 1.71*** | 2.01*** |
| | [1.50,2.01] | [1.27,1.91] | [1.30,1.83] | [1.19,2.22] | [1.41,1.96] | [1.38,2.13] | [1.58,2.54] |
| Global South × Female respondent | | 1.24 | | | | | |
| | | [0.95,1.64] | | | | | |
| Global South × Respondent ≥ 45 | | | 1.34* | | | | |
| | | | [1.00,1.78] | | | | |
| Global South × Higher educated respondent | | | | 1.09 | | | |
| | | | | [0.77,1.53] | | | |
| Global South × High-risk respondent | | | | | 1.18 | | |
| | | | | | [0.86,1.62] | | |
| Global South × High perceived threat respondent | | | | | | 1.03 | |
| | | | | | | [0.78,1.36] | |
| Global South × Employed respondent | | | | | | | 0.80 |
| | | | | | | | [0.60,1.07] |
| Pseudo $R^2$ | 0.10 | 0.10 | 0.10 | 0.10 | 0.10 | 0.10 | 0.10 |
| Observations | 12272 | 11888 | 11888 | 11888 | 11888 | 11888 | 11888 |

**Notes:** Outcome: Choosing the respective candidate to receive the vaccine. Coefficients are Odd's ratios based on conditional logit estimations with standard errors clustered at the individual level. Estimations were conducted with controlling for the main effects of the other three attributes, but only the results for the country of residence attribute are shown here. Columns 2-7 indicate the degree of statistical (in-)significance of the subgroup differences presented in Figure 1 in the main body of the paper and Table A1 of the supplementary material. Results to be interpreted relative to the indicated reference category, i.e., in the case of country of residence, relative to the preference for the vaccine being given to a person living in the country of the survey respondent answering the question. 95% confidence intervals in brackets. * $p < 0.05$, ** $p < 0.01$, *** $p < 0.001$.

**Table S7** – Country of residence attribute: Heterogeneity by respondent's characteristics (French sample)

| | (1) | (2) | (3) | (4) | (5) | (6) | (7) |
|---|---|---|---|---|---|---|---|
| ***Country of Residence*** | | | | | | | |
| Respondents' Country | | | | Reference Category | | | |
| Global South | 1.37*** [1.18,1.59] | 1.31** [1.07,1.59] | 1.32** [1.10,1.59] | 0.95 [0.62,1.44] | 1.39*** [1.16,1.65] | 1.22* [1.01,1.48] | 1.50** [1.13,1.98] |
| Global South × Female respondent | | 1.11 [0.84,1.47] | | | | | |
| Global South × Respondent ≥ 45 | | | 1.12 [0.84,1.50] | | | | |
| Global South × Higher educated respondent | | | | 1.52 [0.98,2.37] | | | |
| Global South × High-risk respondent | | | | | 0.97 [0.71,1.31] | | |
| Global South × High perceived threat respondent | | | | | | 1.29 [0.97,1.71] | |
| Global South × Employed respondent | | | | | | | 0.89 [0.65,1.23] |
| Pseudo $R^2$ | 0.17 | 0.17 | 0.17 | 0.17 | 0.17 | 0.17 | 0.17 |
| Observations | 12256 | 12256 | 12256 | 12256 | 12256 | 12256 | 12256 |

**Notes:** Outcome: Choosing the respective candidate to receive the vaccine. Coefficients are Odd's ratios based on conditional logit estimations with standard errors clustered at the individual level. Estimations were conducted with controlling for the main effects of the other three attributes, but only the results for the country of residence attribute are shown here. Columns 2-7 indicate the degree of statistical (in-)significance of the subgroup differences presented in Figure 1 in the main body of the paper and Table A1 of the supplementary material. Results to be interpreted relative to the indicated reference category, i.e., in the case of country of residence, relative to the preference for the vaccine being given to a person living in the country of the survey respondent answering the question. 95% confidence intervals in brackets. * $p < 0.05$, ** $p < 0.01$, ***$p < 0.001$.

**Table S8** – Country of residence attribute: Heterogeneity by respondent's characteristics (Polish sample)

| | (1) | (2) | (3) | (4) | (5) | (6) | (7) |
|---|---|---|---|---|---|---|---|
| ***Country of Residence*** | | | | | | | |
| Respondents' Country | | | | Reference Category | | | |
| Global South | 0.99<br>[0.86,1.15] | 0.98<br>[0.79,1.21] | 1.03<br>[0.88,1.20] | 1.29<br>[0.97,1.71] | 0.99<br>[0.83,1.18] | 1.00<br>[0.84,1.18] | 1.00<br>[0.77,1.30] |
| Global South × Female respondent | | 1.03<br>[0.78,1.36] | | | | | |
| Global South × Respondent ≥ 45 | | | 0.86<br>[0.61,1.21] | | | | |
| Global South × Higher educated respondent | | | | 0.72*<br>[0.52,0.99] | | | |
| Global South × High-risk respondent | | | | | 1.01<br>[0.76,1.34] | | |
| Global South × High perceived threat respondent | | | | | | 0.99<br>[0.74,1.34] | |
| Global South × Employed respondent | | | | | | | 0.99<br>[0.73,1.34] |
| Pseudo $R^2$ | 0.08 | 0.08 | 0.08 | 0.08 | 0.08 | 0.08 | 0.08 |
| Observations | 10720 | 10720 | 10720 | 10720 | 10720 | 10720 | 10720 |

**Notes:** Outcome: Choosing the respective candidate to receive the vaccine. Coefficients are Odd's ratios based on conditional logit estimations with standard errors clustered at the individual level. Estimations were conducted with controlling for the main effects of the other three attributes, but only the results for the country of residence attribute are shown here. Columns 2-7 indicate the degree of statistical (in-)significance of the subgroup differences presented in Figure 1 in the main body of the paper and Table A1 of the supplementary material. Results to be interpreted relative to the indicated reference category, i.e., in the case of country of residence, relative to the preference for the vaccine being given to a person living in the country of the survey respondent answering the question. 95% confidence intervals in brackets. * $p < 0.05$, ** $p < 0.01$, *** $p < 0.001$.

**Table S9** – Country of residence attribute: Heterogeneity by respondent's characteristics (Swedish sample)

| | (1) | (2) | (3) | (4) | (5) | (6) | (7) |
|---|---|---|---|---|---|---|---|
| ***Country of Residence*** | | | | | | | |
| Respondents' Country | | | | Reference Category | | | |
| Global South | 1.43*** | 1.27* | 1.78*** | 1.39** | 1.60*** | 1.38** | 1.63*** |
| | [1.24,1.65] | [1.02,1.56] | [1.52,2.09] | [1.09,1.76] | [1.37,1.88] | [1.13,1.68] | [1.22,2.17] |
| Global South × Female respondent | | 1.28 | | | | | |
| | | [0.98,1.67] | | | | | |
| Global South × Respondent ≥ 45 | | | 0.48*** | | | | |
| | | | [0.35,0.66] | | | | |
| Global South × Higher educated respondent | | | | 1.05 | | | |
| | | | | [0.79,1.39] | | | |
| Global South × High-risk respondent | | | | | 0.57** | | |
| | | | | | [0.41,0.80] | | |
| Global South × High perceived threat respondent | | | | | | 1.07 | |
| | | | | | | [0.82,1.40] | |
| Global South × Employed respondent | | | | | | | 0.85 |
| | | | | | | | [0.61,1.18] |
| Pseudo $R^2$ | 0.23 | 0.23 | 0.23 | 0.23 | 0.23 | 0.23 | 0.23 |
| Observations | 15008 | 15008 | 15008 | 15008 | 15008 | 15008 | 15008 |

**Notes:** Outcome: Choosing the respective candidate to receive the vaccine. Coefficients are Odd's ratios based on conditional logit estimations with standard errors clustered at the individual level. Estimations were conducted with controlling for the main effects of the other three attributes, but only the results for the country of residence attribute are shown here. Columns 2-7 indicate the degree of statistical (in-)significance of the subgroup differences presented in Figure 1 in the main body of the paper and Table A1 of the supplementary material. Results to be interpreted relative to the indicated reference category, i.e., in the case of country of residence, relative to the preference for the vaccine being given to a person living in the country of the survey respondent answering the question. 95% confidence intervals in brackets. * $p < 0.05$, ** $p < 0.01$, *** $p < 0.001$.

**Table S10** – Country level differences in vaccination rates, willingness and threat perception

| | Germany | Spain | Italy | France | Poland | Sweden |
|---|---|---|---|---|---|---|
| **Time of data collection** | 9.-30.4.21 | 15.-21.6.21 | 15.-21.6.21 | 15.-21.6.21 | 15.-21.6.21 | 15.-24.6.21 |
| **Vaccination rate (first shot)** | 25.3% | 59.9% | 62.3% | 60.2% | 52.5% | 56.8% |
| **Vaccination rate (both shots)** | 8.0% | 36.5% | 31.4% | 30.9% | 36.3% | 34.2% |
| **Vaccination willingness** (control)<br>(1=unsure; 2=depends on vaccine; 3=sure) | 2.14<br>(0.69) | 2.63<br>(0.68) | 2.18<br>(0.77) | 1.77<br>(0.73) | 1.68<br>(0.73) | 2.28<br>(0.81) |
| **Vaccination willingness** (full sample)<br>(1=unsure; 2=depends on vaccine; 3=sure) | 2.22<br>(0.70) | 2.59<br>(0.69) | 2.15<br>(0.76) | 1.79<br>(0.76) | 1.71<br>(0.76) | 2.29<br>(0.80) |
| **COVID-19 threat perception** (initial coding)<br>(Germany: 7-point scale; Other countries: 5-point scale) | 4.57<br>(1.67) | 3.52<br>(1.12) | 3.71<br>(1.06) | 3.36<br>(1.10) | 2.97<br>(1.20) | 3.39<br>(1.08) |
| **Elevated COVID-19 threat perception**<br>(dummy coded, Germany: 1-4=0; 5-7=1; Other countries: 1-3=0; 4-5=1) | 0.54<br>(0.50) | 0.53<br>(0.50) | 0.58<br>(0.49) | 0.46<br>(0.50) | 0.33<br>(0.47) | 0.49<br>(0.50) |

**Notes:** The vaccination rates reported here are the averages for the time during which each survey was in the field. Data was drawn from https://vaccinetracker.ecdc. europa.eu/public/extensions/COVID-19/vaccine-tracker.html. The vaccine willingness results were calculated both for the full sample and for a smaller subsample. The latter served as the control group in another survey experiment conducted throughout this same data collection, in which participants received different messages intended to reduce vaccine hesitancy. Thus, to make sure the vaccine willingness results are unaffected by this, we additionally report values from the control group.

### Figures

**Figure S1** – Timeline survey launch and infection rates

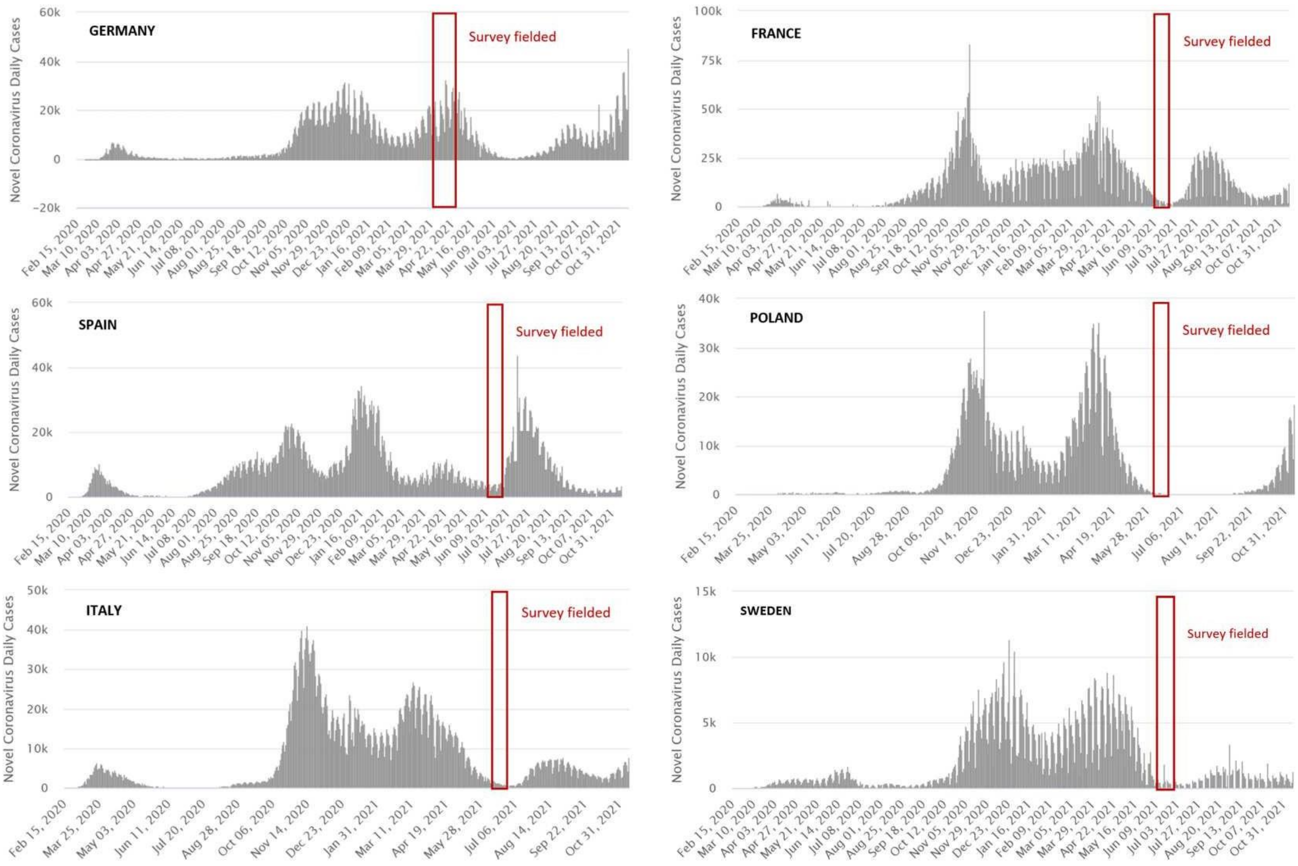


Notes: Source: https://www.worldometers.info/coronavirus

**Figure S2** – Sample choice set presented to respondents

**Which of the following two persons should receive the vaccine first, person A or person B?**

| | **Person A** | **Person B** |
|---|---|---|
| **Age** | 20 years old | 40 years old |
| **Risk of COVID-19 death** | Increased risk due to comorbidity and/or lifestyle | No increased risk due to comorbidity and/or lifestyle |
| **Employment status** | Employed in essential services (e.g. health personel, supermarket employee) | Employed, and income losses due to COVID-19 restrictions |
| **Country of residence and health care system capacity** | Low-income country, with low healthcare system capacity (e.g. India, Nigeria, Bolivia) | Germany, with high healthcare system capacity |

| | Person A | Person B |
|---|---|---|
| **Your decision:** | ○ | ○ |

*Technical notes: The design was determined to be D-efficient based on weak priors for the main attributes effects (without interactions). Statistical efficiency was measured by the D-optimality criterion (D-error), the most widely used metric in this regard. D-optimal or D-efficient designs minimise the determinant of the asymptotic variance-covariance matrix, ensuring minimum variation around the parameter estimates.*

**Figure S3** – Country of residence attribute: Heterogeneity by respondent's characteristics(by country)

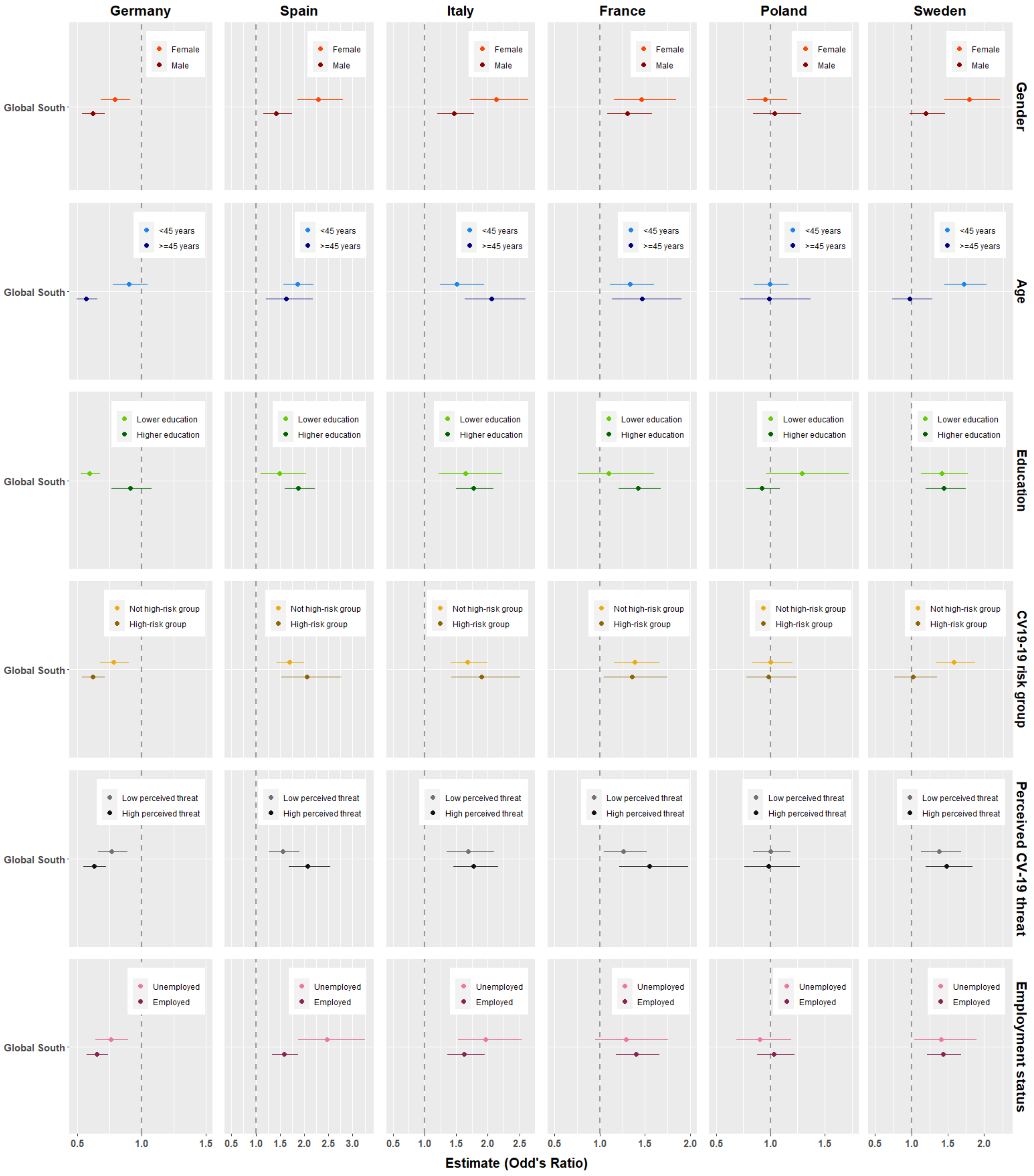

**Figure S4** – COVID-19 threat perception across countries

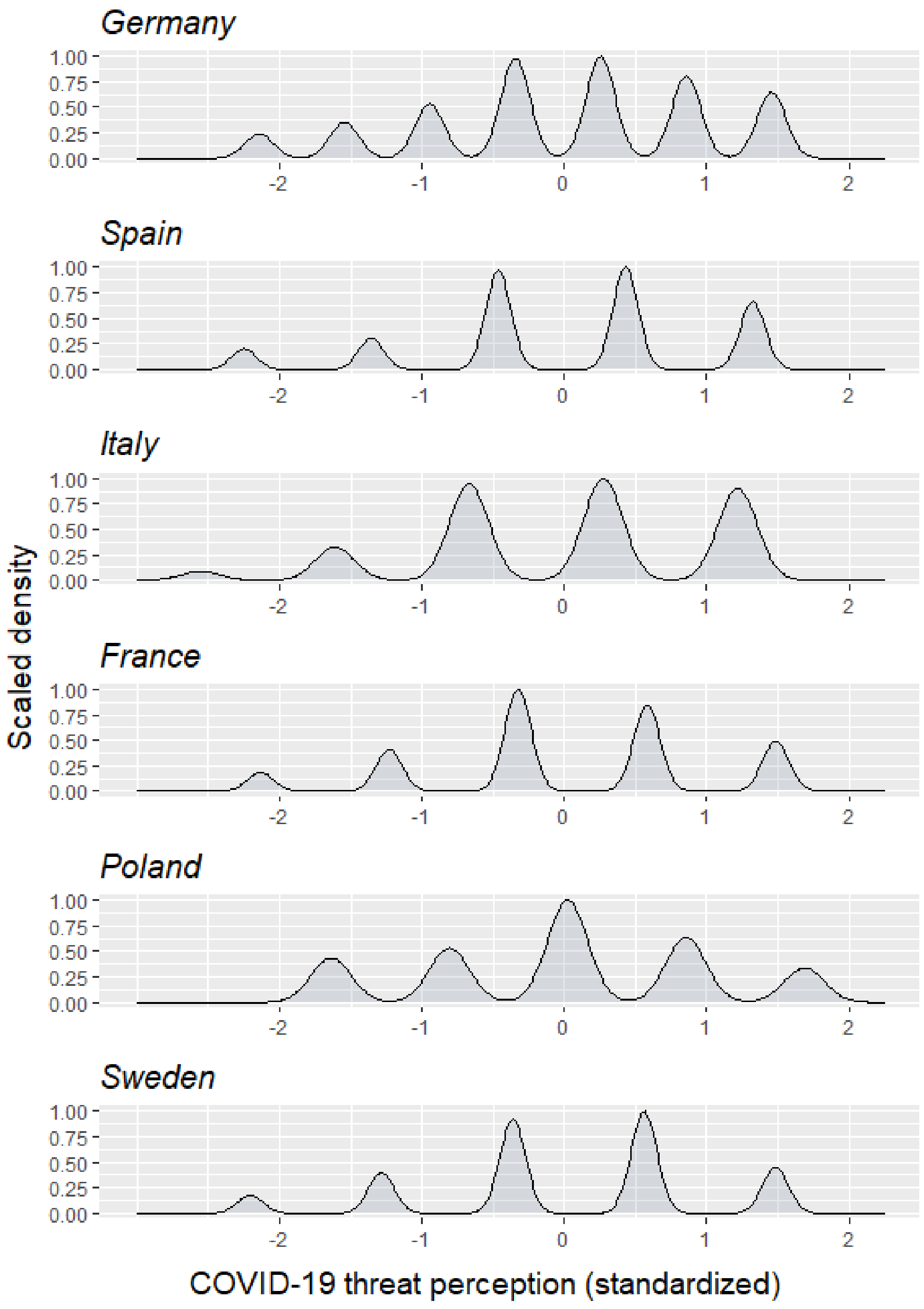
